# Perioperative changes in oxygen consumption: estimations from minimal-invasive cardiac output and a-cvO_2_ difference parallel to indirect calorimetry

**DOI:** 10.1101/2022.07.18.22277772

**Authors:** Julia Jakobsson, Carl Norén, Eva Hagel, Magnus Backheden, Sigridur Kalman, Erzsébet Bartha

## Abstract

Feasible estimations of perioperative changes in oxygen consumption could enable larger studies of its role in postoperative outcomes. Current methods, by pulmonary artery catheterisation or breathing gas analysis, are often regarded as either invasive or technically requiring. In this pilot study, we investigated the relationship between estimations of oxygen consumption, based on minimal-invasive cardiac output and arterial-central venous blood gas sampling, and indirect calorimetry in the perioperative period using the data collected during a clinical trial on perioperative oxygen transport.

In 20 patients >65 years during epidural and general anaesthesia for open abdominal surgery, Fick-based estimations of oxygen consumption(EVO2), the product of cardiac output from LiDCO™plus(LiDCO Ltd, Cambridge, UK) and arterial-central venous oxygen content difference, were compared with indirect calorimetry(GVO_2_) using QuarkRMR(COSMED, srl. Italy). Eighty-five simultaneous intra- and postoperative measurements at different time-points were analysed for prediction, parallelity and by traditional agreement assessment. There was an overall association between GVO_2_ and EVO_2_, 73(95% CI 62 to 83) + 0.45(95% CI 0.29 to 0.61) EVO_2_ ml min^-1^m^-2^, *P*<0.0001. GVO_2_ and EVO_2_ changed in parallel intra- and postoperatively when normalised to their respective overall means. Unadjusted mean difference between GVO_2_ and EVO_2_ indexed for body surface area was 26(95% CI 20 to 32) with limits of agreement (1.96SD) of -32 to 85 ml min^-1^m^-2^ and did not change over time. There was low correlation for absolute agreement, ICC(A,1) 0.37(95% CI 0.34 to 0.65) [F(84,10.2)=3.07, *P*=0.0266].

Despite lack of absolute agreement, the estimated oxygen consumption changed in parallel to the metabolic measurements in the perioperative period. Prediction or trending of oxygen consumption by this or similar methods could be further evaluated in larger samples.

## Introduction

A postoperative imbalance between oxygen consumption and delivery, leading to increased oxygen extraction, has been associated with increased morbidity and mortality after major surgery.(1) The focus of goal-directed haemodynamic therapy (GDHT) has traditionally been on oxygen delivery, which is often easier to assess and to develop measurable optimisation strategies for.(2) Recently, interest is growing to reassess perioperative oxygen consumption in current surgical populations using modern monitoring and analytic methodologies.(3-5) Feasible estimations could enable larger studies on the role of oxygen consumption in postoperative outcomes. Available techniques, by pulmonary artery catheterisation or indirect calorimetry, are either deemed too invasive or difficult to manage in a clinical study setting during non-cardiac surgery. Using oxygen uptake calculated from fractions of inspiratory and expiratory oxygen in the closed breathing circuit during low-flow anaesthesia(6) has not demonstrated agreement when compared to standard methods.(7) Importantly, it can not be used in awake patients in the postoperative period. Commonly used haemodynamic monitoring in major surgery, such as minimal-invasive cardiac output with arterial and central venous access, could offer a possibility not only to estimate intra- and postoperative oxygen consumption but also to follow changes over time. By substituting mixed with central venous oxygen content and using the cardiac output derived from a minimal-invasive monitor, an estimation of oxygen consumption can be calculated by the reverse Fick principle.(8) The lack of absolute agreement between calorimetric and Fick-based methods has been reported previously, the latter do not include pulmonary oxygen consumption and global oxygen consumption values are usually reported around 20-40 ml min^-1^m^-2^ lower compared to those obtained from breathing gas analysis.(9-11) Yet, if this bias remains unchanged in the intra- and postoperative period, such estimations could be studied in larger samples and related to other clinical parameters and outcomes.

Our aim of this pilot study was to investigate the relationship and temporal changes between estimations of oxygen consumption, based on minimal-invasive cardiac output monitoring and arterial-central venous blood gas samples (EVO_2_), and measurements by indirect calorimetry (GVO_2_) in the perioperative period using the data collected during our oxygen transport study in elderly undergoing major abdominal surgery.(12)

## Materials and methods

The present analysis was a secondary objective of a prospective observational study on perioperative oxygen transport in elderly patients undergoing major upper abdominal surgery (clinicaltrials.gov NCT03355118). The Regional Ethics Review Board of the Stockholm Region (ID 2017/291-31/4) approved the study and written informed consent was obtained from all participants. The primary aim of the study, i.e. the perioperative oxygen transport changes, has been published.(12) Data collection and analysis for the present publication were pre-planned and conducted simultaneously. The original cohort data was prospectively collected 2017-2018.

### Patients and settings

A detailed description of the selection criteria, patient characteristics, perioperative management and oxygen transport outcomes can be found in the previous publication.(12) As stated there, 20 ASA II-IV patients over 65 years undergoing open pancreatic or liver resection surgery in epidural and general anaesthesia were included. The study was conducted at the Karolinska University Hospital in Huddinge, a tertiary referral center for upper abdominal surgery.

### Data extraction and time-points

Paired values of oxygen consumption by estimations based on minimal-invasive cardiac output monitoring and arterial-central venous blood gas samples (EVO_2_) and indirect calorimetry GVO_2_) from five perioperative time-points were analysed; T1: during anaesthesia, right before surgical skin incision; T2: early during surgery, directly after skin incision; T3: later during surgery, >2h after skin incision; T4: early postoperatively, <12h after extubation; T5: late postoperatively, on postoperative day 1. The mean values for GVO_2_ during the approximate 20-minute measurement periods were compared with simultaneous cardiac output measurements averaged for each minute exported from LiDCOviewPRO (LiDCO Ltd, Cambridge, UK). The blood gas parameters were calculated as means of two simultaneously drawn arterial and central venous samples at 5 and 15 minutes into the measurement period.

### Measurements of VO2 by indirect calorimetry (GVO2)

Indirect calorimetry was performed by QuarkRMR (COSMED srl, Italy). This device applies a breath-by-breath technique to measure gas flow and concentrations that are synchronised by data processing algorithms. The Haldane transformation is used to calculate oxygen consumption.(13) During intraoperative measurements, the flow meter (Flow-REE, COSMED srl, Italy), gas sampling line and moist filter were placed between the endotracheal tube and the Y-piece of the ventilator. The ventilator was set to a fresh gas flow of 2 L min^-1^ and FiO_2_ of 0.5 during measurements to allow for gas sampling. All other ventilation settings were left unchanged. Postoperative measurements were made with a tight-fitting face mask connected to a bidirectional turbine flow meter and a gas sampling line. No supplemental oxygen was administered during the postoperative measurements. The calorimeter was calibrated before start of intraoperative measurements and before each postoperative measurement after a warm-up time of 20 minutes with a standardised gas mixture containing 16% oxygen and 5% carbon dioxide. The gas sampling line, Flow-REE and moist filter were changed before each measurement (except before T2, continuous to T1) and all flowmeters were calibrated with a 3L-syringe.

### Estimation of VO2 by minimal-invasive cardiac output and arterial-central venous oxygen content difference (EVO2)

EVO_2_ was calculated by the reverse Fick’s principle with central venous instead of pulmonary artery blood using the following formulas:(14)

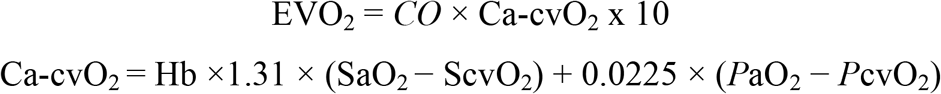

[*CO*; cardiac output in L min^-1^, Ca-cvO_2_; oxygen content difference between arterial and central venous blood in ml dl^-1^, Hb; haemoglobin in g dl^-1^, SaO_2_; arterial oxygen saturation, ScvO_2_; central venous saturation, *P*aO_2_; partial pressure of oxygen in arterial blood, *P*cvO_2_; partial pressure of oxygen in central venous blood, constants 1.31 and 0.0225, referring to the Hüfner constant and the solubility coefficient of oxygen (ml O_2_ dl^-1^ kPa^-1^), and 10 as a conversion factor from dL to L.]

Cardiac output was obtained from LiDCO™plus (LiDCO Ltd, Cambridge, UK). The device was calibrated and recalibrated according to the manufacturer’s instructions. Missing values from *CO* measurements (averaged for each minute) were substituted by linear interpolation if no more than 3 data points and no major haemodynamic changes occurred. Blood gases were analysed immediately after sampling by ABL800 Flex or ABL90 Flex (Radiometer Medical ApS, Denmark). Cardiac output and measured oxygen consumption were indexed for body surface area using the DuBois formula yielding values of GVO_2_ and EVO_2_ in ml min^-1^m^-2^.(15)

### Statistical analysis

The sample size calculation was performed for the primary study, in which 20 patients were expected to demonstrate a relevant change in oxygen consumption after induction of anaesthesia. This would yield a maximum of 100 paired measurements of EVO_2_ and GVO_2_ which was considered sufficient even in the presence of >10% data loss. Continuous data was tested for normality distribution and statistical tests applied accordingly. Statistical analyses were performed and constructed in R (version 3.5.3; R Foundation for Statistical Computing, Vienna, Austria, URL; https://www.R-project.org) and SAS (version 9.4; SAS Institute Inc, Cary, NC, U.S.). The statisticians conducting the analyses were not involved in the data collection. Mean difference between EVO_2_ and GVO_2_ with 95% confidence interval were calculated from the individual paired measurements and grouped by time point (T1-5). These changes over time were analysed by linear mixed models with Holm-adjusted Tukey post-hoc tests. To investigate the overall association between EVO_2_ and GVO_2_, a random coefficient model was used based on individual slopes and coefficients. Analyses of the perioperative changes over time of GVO_2_ compared to EVO_2_ and its input variables (*CI*; cardiac index and Ca-cvO_2_) were conducted by random effect mixed models with method or component and time as fixed effects. Adjustment for differences in variances of the methods or components was made. In these models, the relative changes were normalised to the patients’ individual baseline measurements (T1). In the model analysing changes of each method in awake and anaesthetised subjects, the changes were normalised to the respective overall mean. Traditional agreement assessment was also performed by intraclass correlation and Bland-Altman analysis. Single score intraclass correlation was used, a in a two-way model yielding ICC coefficients with 95% CI. Bias and limits of agreement with 95% CI was visualised in Bland-Altman plots. Both ICC and Bland-Altman analyses were performed separately for each time-point T1-T5. The overall ICC and Bland-Altman analyses were not adjusted for repeated measurements as these were performed under varying intra- and postoperative conditions. Normality and homoscedasticity were assessed in residual plots. An alpha of 0.05 was considered significant.

## Results

A total of 85 paired measurements of EVO_2_ and GVO_2_ were obtained in 20 subjects; 58 were obtained intraoperatively and 27 in the postoperative period. Four paired intraoperative measurements were not performed due to early termination of surgery (unexpected metastatic spread of malignancy) in two patients. Thirteen paired measurements could not be performed in the postoperative period because of technical or arterial line failure (n=2), logistical reasons (n=2), patients’ decline (n=3), exclusion due to short postoperative stay (n=4) and need for supplemental oxygen (n=2). Correct positioning of the CVC was confirmed by postoperative chest x-ray in all patients.

There was a overall mean difference between EVO_2_ and GVO_2_ [F(1, 167) = 72.8, *P*<0.0001] estimated to -26 (95% CI -20 to -32; *P*<0.001) ml min^-1^m^-2^. This difference was largely unchanged at the different time-points [F(4, 168)=1.39, *P*=0.241]. The means of GVO_2_ and EVO_2_ at the different perioperative time-points (T1-T5) are presented in Fig 1.

**Fig 1.**
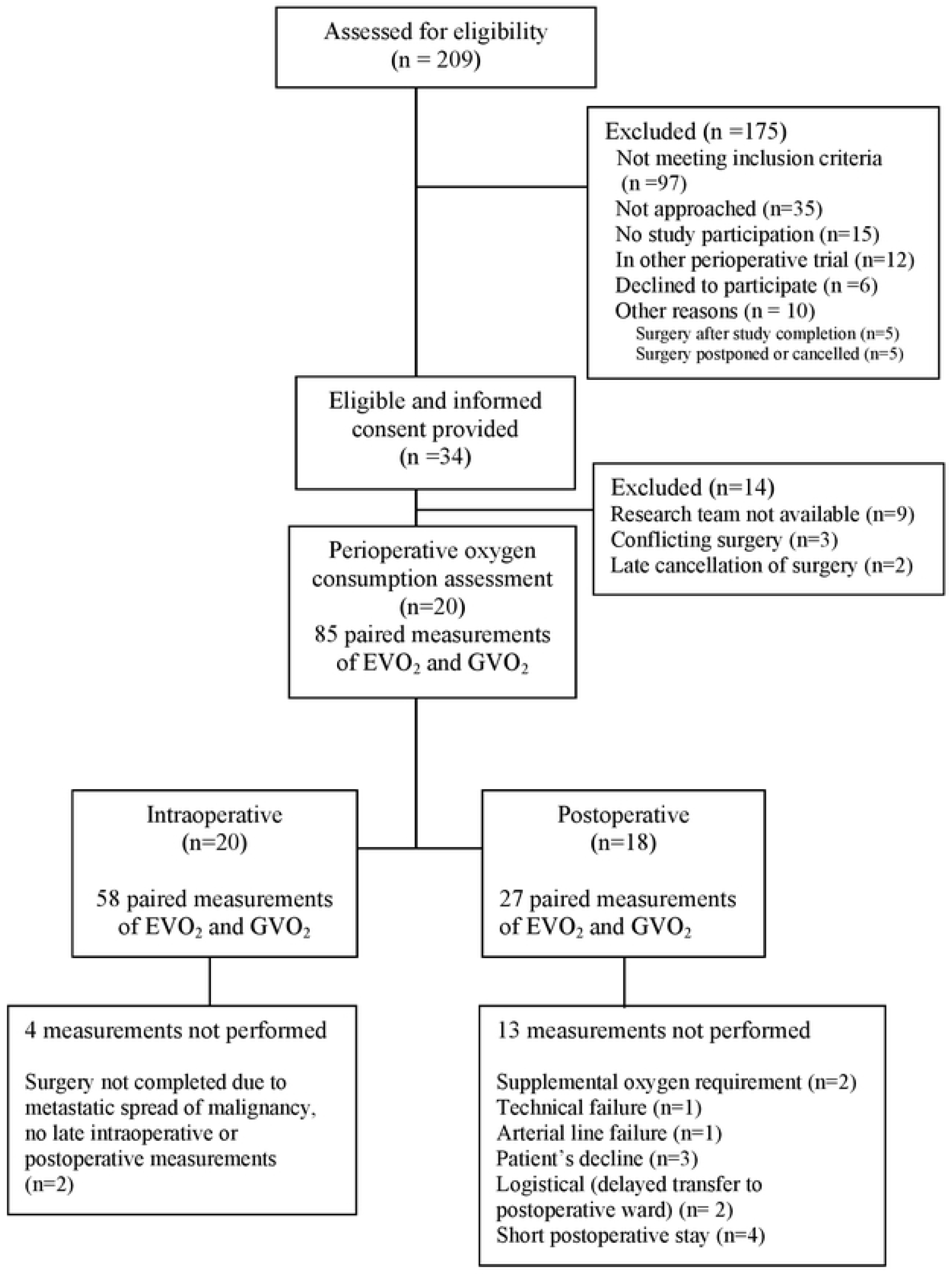
CONSORT flow diagram. Enrollment and assessment of estimated (EVO_2_) and measured (GVO_2_) oxygen consumption (n) patients.

GVO_2_ and EVO_2_ changed in parallel when separated to the anaesthetised intraoperative state [F(2, 49.9)=0.57, *P*=0.5669] and the awake postoperative state F(1, 22) = 0.00, *P*=0.9604), see Figs 2a-b. An overall association between GVO_2_ and EVO_2_ was demonstrated in a random coefficient model (Fig 3). The two patients with early termination of surgery were excluded from this analysis. The variances of EVO_2_ and its components, oxygen content difference in arterial and central venous blood (Ca-cvO_2_) and cardiac index (*CI*) were larger compared to GVO_2_ at all time-points, these analyses are presented in Suppl S1.

**Fig 2.**
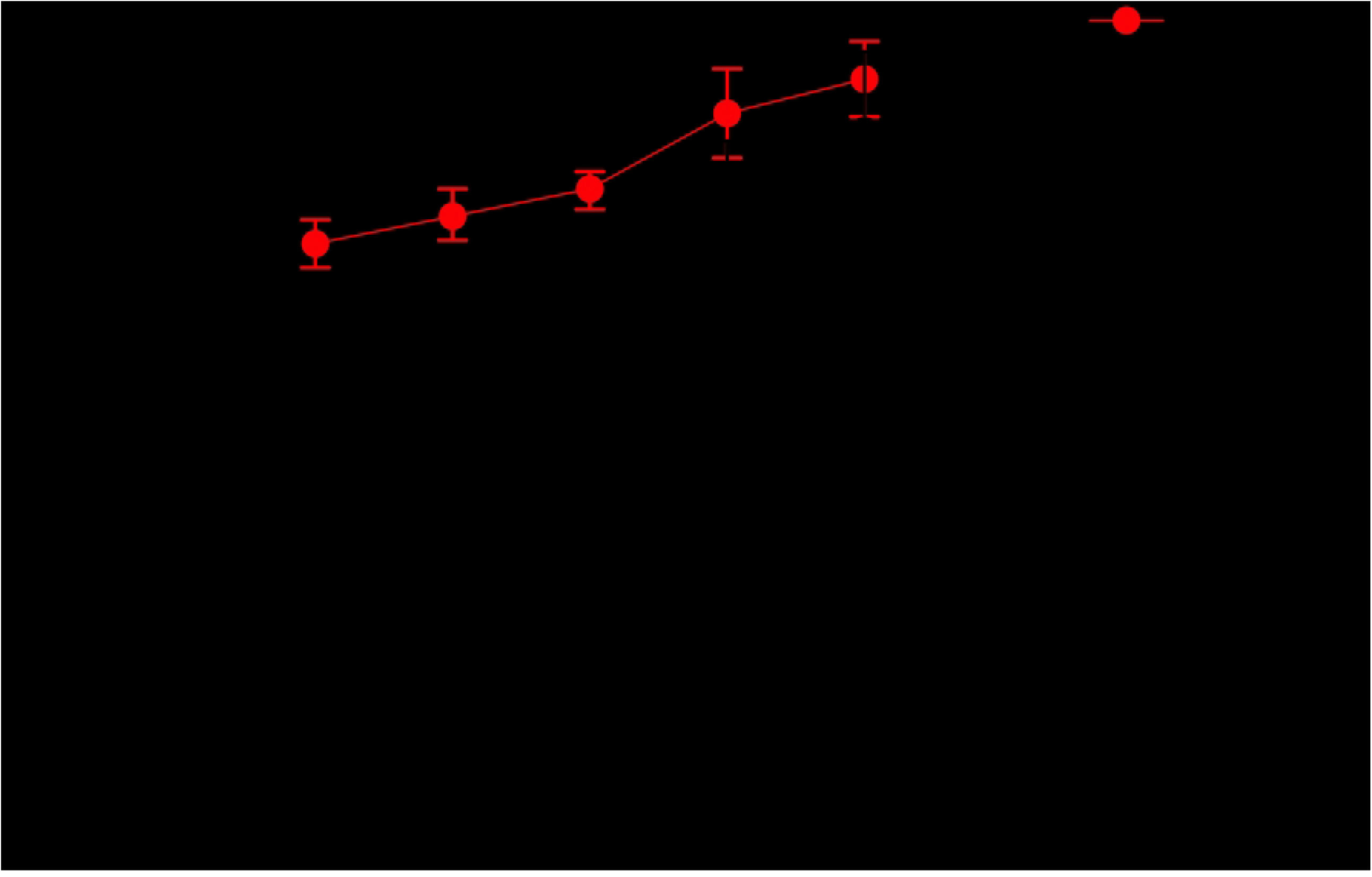
Oxygen consumption (VO_2_) at each perioperative time-point (T1-5). EVO_2_ (estimated from minimal-invasive cardiac output and arterial-central venous blood sampling) and GVO_2_ (indirect calorimetry) expressed as mean (95% CI) ml min^-1^ m^-2^.

**Figs 3a-b.**
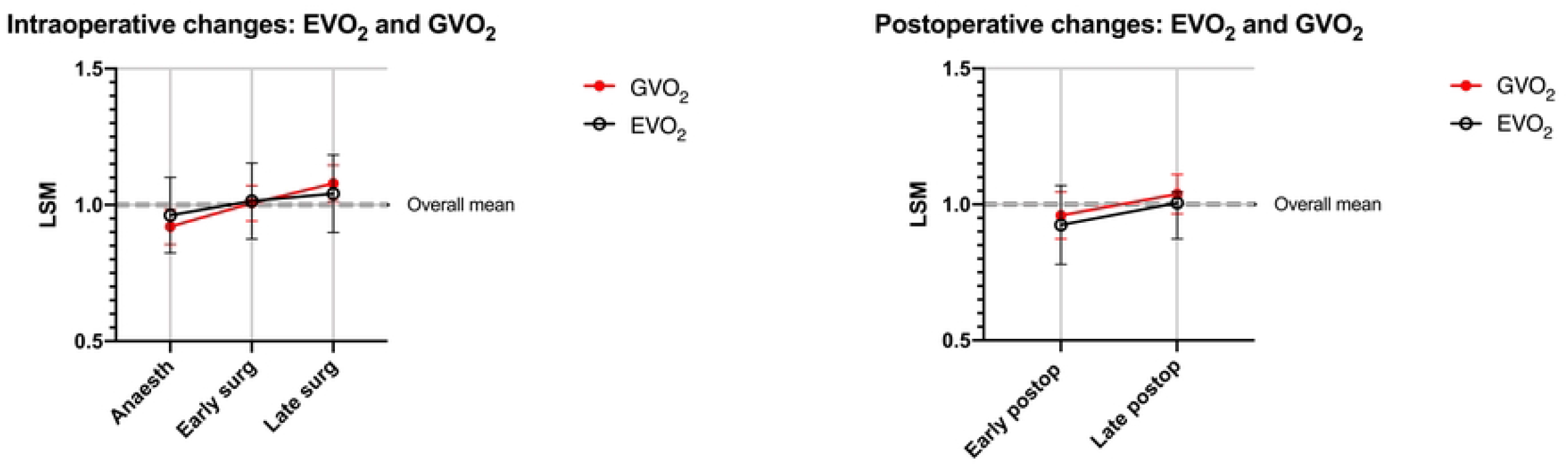
Results from the mixed effect models on perioperative changes of GVO_2_ (red) and EVO_2_ (black). Least square means estimates with 95% CI and normalised to overall means (=1.0) of each method in anaesthetised intraoperative (T1-T3) and awake postoperative states (T4-T5). (T1) anaesthesia; Early surgery (T2); Late surgery (T3); Early postop (T4); Late postop (T5).

**Fig 4.**
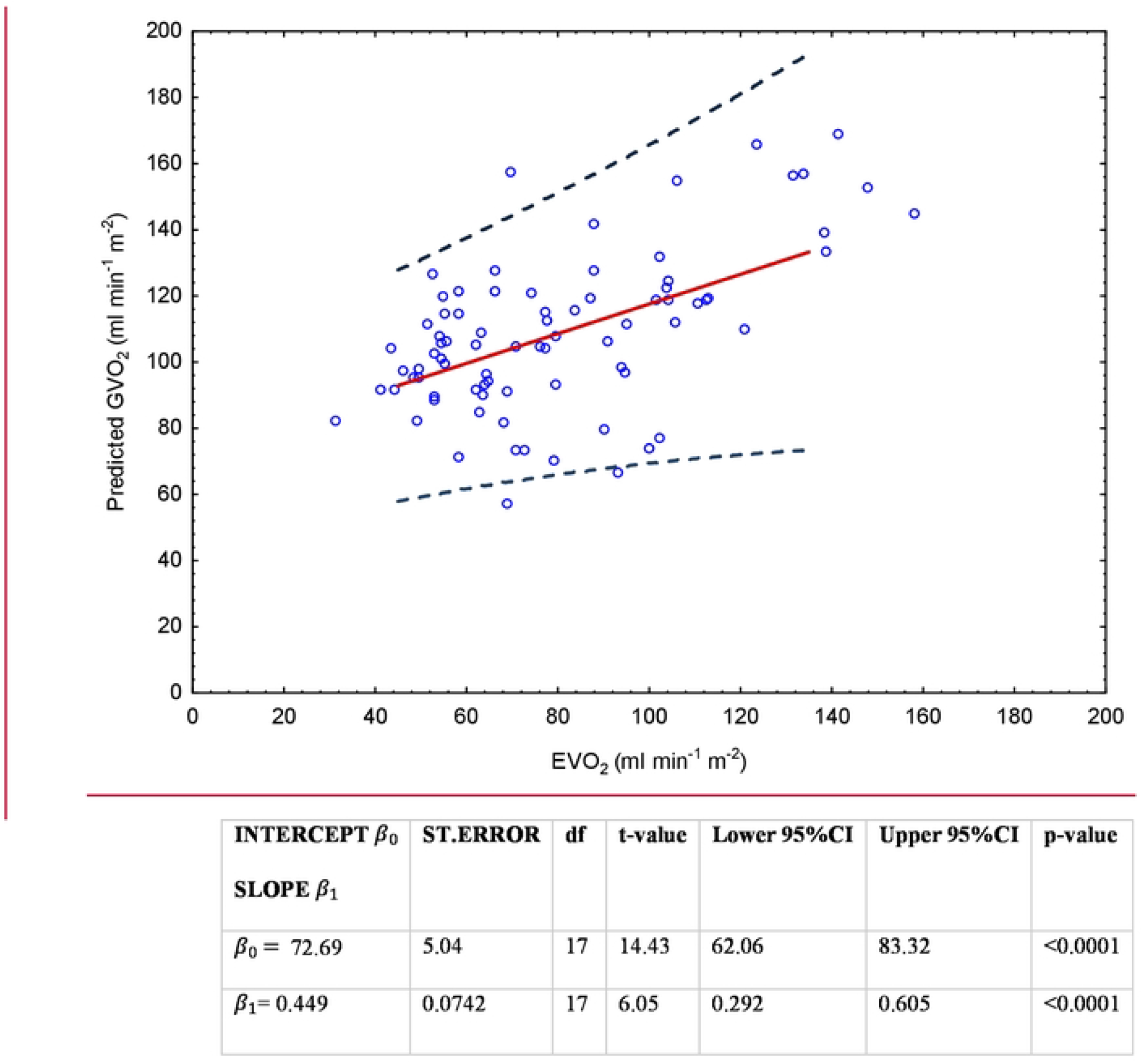
A random coefficient model for predicted GVO_2_ from EVO_2_ based on all perioperative time-points. GVO_2_ = β_0_ + β_1_ (EVO_2_).

The traditional agreement analyses are presented as supplementary results. The overall unadjusted mean bias was 26 ml min^-1^ m^-2^ with limits of agreement (1.96SD) of -32 to 85 ml min^-1^m^-2^. Excluding one outlier in the late postoperative period (a patient with a large Ca-cvO_2_ difference) changed the unadjusted bias to 28 (LoA -20 to 75) ml min^-1^ m^-2^. Bland-Altman plots were constructed to illustrate the bias and limits of agreement at the different time-points Suppl S2. The overall correlation for *absolute* agreement was poor, with an intraclass coefficient ICC(A,1) of 0.37 (95% CI 0.34 to 0.65) [F(84,10.2)=3.07, *P*=0.0266], and did not improve much when adjusted for lower overall mean difference of EVO_2_, ICC(A,1)=0.51 (95% CI 0.34 to 0.65) [F(84, 84) = 3.07, *P*<0.001]. Graphs depicting the correlation between indexed GVO_2_ and EVO_2_ at the different time-points (T1-5) including the unadjusted overall correlation are presented in Suppl S3.

## Discussion

Estimates of increased oxygen extraction, i.e low mixed or central venous saturation, are associated with poor surgical outcomes.(16, 17) However, cut-off levels remain unclear and the quality of evidence is low.(18) In order to further study and distinguish the role of oxygen consumption in the perioperative period, feasible estimations are needed. To the best of our knowledge, this is the first pilot study investigating how a Fick-derived estimation method based on minimal-invasive haemodynamic monitoring (LiDCO™plus and blood gas sampling from arterial and central venous lines) can be used intra- and postoperatively. As expected, this estimation method had poor agreement concerning absolute values when compared to indirect calorimetry, but approximate to previous studies using pulmonary artery catheters.(11, 19-21) Importantly, the estimations based on routine haemodynamic monitoring for major surgery, were shown to change in parallel with the metabolic measurements in the perioperative period. We suggest that this or similar methods could be evaluated in larger samples and related to clinical outcomes.

Most previous studies investigating methods for oxygen consumption monitoring perioperatively or in critically ill patients were performed decades ago using traditional method comparison analytical methods. Some of the earlier method comparison studies are summarised in Table 1. Newer studies using non-invasive cardiac output monitors have not shown agreement with oxygen consumption measurements from indirect calorimetry(22) or pulmonary artery catheters.(23) However, the monitors used were not calibrated by transpulmonary or indicator dilution such as the PiCCO™ or LiDCO™plus systems and did not analyse changes over time.

**Table 1.** Examples of previous studies comparing methods for gas-derived VO2 with Fick-derived VO2. Abbreviations: IC; indirect calorimetry: PAC; pulmonary artery catheter: B-A; Bland-Altman method for assessing agreement: ANOVA; analysis of variance; PE: pure error; RE: relative error: ICU; intensive care unit: CC; closed circuit anaesthesia system: CVC; central venous catheter: PDD; pulse dye densitometry: * no overall LoA. ** derived from graph ***pre-CPB measurements

| Author, year | Subjects, N= | Number of paired measurements | Method derived $\text{VO}_2$ gas- | Method derived $\text{VO}_2$ Fick- | Bias (SD or 95% CI), limits of agreement in $\text{ml min}^{-1} \text{m}^{-2}$ | Statistical methodology |
| --- | --- | --- | --- | --- | --- | --- |
| Bizouarn et al.<sup>16</sup>, 1992 | Postop cardiac surgery, N=10 | 50 | IC Deltatrac® | PAC thermodilution | 34 (SD 27)<br>LoA: -33 to 88 | B-A<br>ANOVA for time-effects |
| Bizouarn et al.<sup>22</sup>, 1995 | Postop cardiac surgery, N=9 | 54 | IC Deltatrac® | PAC (continuous thermodilution) | 15 (95% CI, 13 to 17)<br>LoA: -3 to 33 | B-A<br>PE-RE |
| Epstein et al.<sup>12</sup>, 2000 | Trauma ICU, N=38 | 152 | IC Puritan Bennett® | PAC thermodilution | 41 (95% CI, 20 to 63)<br>LoA*: lower -40 to -72, upper 120 to 149 | B-A |
| Hofland et al.<sup>14</sup>, 2003 | Intraop vascular surgery, N=11 | 73 | CC Physioflex® | PAC thermodilution | 36 (not presented)<br>LoA**: -40 to 112 | B-A<br>Linear regression/Spearman rank correlation |
| Inadomi et al.<sup>5</sup>, 2008 | Postop major abdominal surgery, N=28 | 56 | IC Puritan Bennett® | CVC+PDD | 33 (not presented)<br>LoA: -31 to 97 | B-A<br>Linear regression |
| Keinanen and Takala<sup>7</sup>, 1997 | Periop cardiac surgery, N=9 | 45 | IC Deltatrac® | PAC thermodilution | 33 (25)<br>LoA (not analysed) | Linear regression<br>ANOVA |
| Leonard et al.<sup>4</sup>, 2002 | Periop cardiac surgery, N=29 | 29*** | CC Biro method | PAC thermodilution | 75 (121)<br>LoA: -162 to 311 | B-A |
| Myles et al.<sup>17</sup> 1996/revised<sup>15</sup> 2007 | Periop cardiac surgery, N=20 | 143 | IC Deltatrac® | PAC thermodilution | 20 (50)<br>LoA -128 to 88, revised 30 (-116 to 57) | B-A (1996)<br>Random effects model (2007) |
| Peyton and Robinson<sup>8</sup>, 2005 | Intraop cardiac surgery, N=9 | 18 | Modified Bains circuit | PAC thermodilution | 19 (20) (95% CI, 9 to 29) $\text{ml min}^{-1}$<br>LoA (not analysed) | Mean difference |
| Saito et al.<sup>9</sup>, 2007 | Periop oesophagectomy, N=35 | 210 | IC Deltatrac® | PAC thermodilution | 23 (95% CI, 20 to 27)<br>LoA: -23 to 69 | B-A<br>Correlation<br>Difference over time |
| Smithies et al.<sup>23</sup>, 1991 | General ICU, N=8 | 20 | CC spirometry | PAC thermodilution | 36 (SD29) $\text{ml min}^{-1}$<br>LoA (not analysed) | Mean difference |
| Soussi et al.<sup>10</sup>, 2017 | ICU burns patients, N=22 | 44 | IC E-COVX® | CVC +PiCCO® | 60 (not presented)<br>LoA: -84 to 203 | Linear regression<br>Bland-Altman |
| Stuart-Andrews et al.<sup>24</sup>, 2007 | Intraop cardiac surgery, N=30 | 30*** | Modified semi-closed breathing circuit | PAC thermodilution | 21 (25)<br>LoA (overall in graph) | Correlation<br>Bland-Altman |
| Walsh et al.<sup>25</sup>, 1998 | ICU hepatic failure, N=17 | 98 | IC Deltatrac® | PAC thermodilution | -41(30) (95 % CI, -31 to -47)<br>LoA: -101 to 19<br>(Fick - Gas) | Bland-Altman<br>Repeatability |

A feasible oxygen consumption estimation method does not necessarily have to demonstrate perfect agreement to reflect changes in the perioperative period. Time effects and repeated measurements in the same subject under changing conditions are important statistical challenges in studies involving perioperative patients. Previous studies have often used simple linear regression or correlation(9, 10, 22, 24) or Bland-Altman analysis(25) without correction for repeated measurements(11, 22, 26, 27) except for some.(21, 28) Only a few addressed the relationship between measurements over time.(19, 29) In the present study, we developed a prediction model for EVO_2_ and GVO_2_ by using a random coefficient model based on individual slopes and intercepts. A significant positive association was demonstrated here, but such prediction models should obviously be evaluated in larger samples. We also present analyses of relative changes of EVO_2_ and its components with GVO_2_. The parallelity that was demonstrated could indicate an ability of EVO_2_ to track changes in oxygen consumption. To address this further, multiple measurements during shorter periods of time would be required. Analytic models previously used for cardiac output monitors such as polar plot approaches could be used to assess the magnitude and direction of changes.(30) Intraclass correlation (ICC) was used as it better reflects reliability and agreement based on analysis of variance of the pooled data.(31) When adjusted for the consistently lower values of EVO_2_, the ICC estimates of the model improved but not so much (ICC coefficient 0.51 vs 0.37). Bland-Altman analysis has since long been the standard method for visualisation of agreement when comparing different methods of VO_2_ monitoring.(21) Myles and Cui further elaborated the methodological issues related to repeated measurements in the same subject already considered by Bland and Altman(32) and proposed different random effects models to adjust limits of agreement.(21) As measurements were performed under varying perioperative conditions, we present the time-points separately and did not adjust the overall limits of agreement for repeated measurements in the same patient.

Oxygen consumption calculated by the reverse Fick equation is consistently reported lower than simultaneous measurements by analysis of respiratory gas exchange.(7, 9-11, 19, 24, 26, 28, 29, 33-36) This difference or bias has been attributed to the pulmonary oxygen consumption.(35, 37, 38) However, variability of Fick-derived measurements(34, 36)and wide limits of agreement(21) has made it difficult to estimate a systematic methodological bias. Many previous studies have either been performed in thoracic or cardiovascular surgery(9-11, 19, 37) or in critically ill patients.(24, 26, 36, 39) Pulmonary VO_2_ can be expected to increase after thoracic surgery(11) and in intensive care patients with varying degrees of lung injury.(40) Some studies that involve patients undergoing predominately abdominal surgery have shown acceptable agreement between the methods.(27, 41)The age of the studies is also reflected by the frequent use of the Deltatrac Metabolic Monitor® (Datex Instrumentarium, Helsinki, Finland), a metabolic monitor using a mixing-chamber technique and which is no longer in production. Many metabolic monitors in modern clinical use are based on breath-by-breath technology such as the Es-COVX® (GE Healthcare, Helsinki, Finland) or the QuarkRMR® in our study. Although there is supporting evidence for some overestimation of VO_2_, the technology has shown clinically acceptable agreement when compared with mixing-chamber methods(42, 43) and it has been validated in the semi-closed circle absorber systems commonly used in anaesthesia.(44) Our results on GVO_2_ were comparable with studies using Deltatrac II when corrected for difference in units (*Table 1*). The estimations of oxygen consumption rely on accurate cardiac output determinations and oxygen content difference measurements. The LiDCO™plus has shown acceptable performance against the pulmonary artery catheter and other devices in cross-comparisons in cardiac output accuracy studies. (45, 46) During rapidly changing haemodynamic situations, concerns regarding trending ability and underestimation of cardiac output have been raised.(30, 47) The 20-minute data extraction periods in this study were specifically chosen to represent perioperative time points that usually are without considerable circulatory instability. Central and mixed venous oxygen saturation have not shown interchangeability(48-50) but some studies have suggested that trends in ScvO_2_ can replace SvO_2_.(51-53) During stable intraoperative conditions, oxygen content difference is not expected to vary to a large extent whereas cardiac output can show considerable in- and between patient variability.(23) In our study, oxygen content difference and cardiac output demonstrated similar coefficients of variation.

The present study has several major limitations in addition to those discussed above. The sample size and number of observations are small, although in the same range as many earlier studies comparing calculated and measured VO_2,_ and this could explain the large variability of EVO_2._ There was a considerable loss of data in the postoperative period limiting the conclusions on changes over time.

In summary, we have evaluated an oxygen consumption method that requires no extra equipment if perioperative CO-monitoring and CVC are used. Our results indicate that the performance could be equivalent to pulmonary artery catheters, and we have demonstrated parallelity in changes over time. Compared to earlier studies, we have used modern and commonly used calibrated CO-monitoring. We have also addressed the relationship between measurements over time using updated statistical analysis. The results should be regarded as indicative and further studies on larger samples are needed to establish if this method of estimating perioperative VO_2_ can prove useful.

## Data Availability

All relevant data are within the manuscript and its Supporting Information files

## Acknowledgements

The authors would like to thank research assistant nurses at the Clinical Research Unit, Dept. of Upper Abdominal Surgery, Karolinska University Hospital Huddinge, Birgitta Holmberg and Sirje Laur, for their help in recruiting and informing the patients included in this study.

## Statements and declarations

The authors have no conflicts of interest to declare.

## Supporting information

**S1 File**

**S2 File**

**S3 File**

## References

1. Shoemaker WC, Appel PL, Kram HB. Hemodynamic and oxygen transport responses in survivors and nonsurvivors of high-risk surgery. Crit Care Med. 1993;21(7):977–90.

2. Grocott MP, Dushianthan A, Hamilton MA, Mythen MG, Harrison D, Rowan K, et al. Perioperative increase in global blood flow to explicit defined goals and outcomes following surgery. Cochrane Database Syst Rev. 2012;11:CD004082.

3. Maheshwari K. Principles for minimizing oxygen debt: can they translate to clinical application and improve outcomes? Best Practice & Research Clinical Anaesthesiology. 2020.

4. Jakobsson J, Vadman S, Hagel E, Kalman S, Bartha E. The effects of general anaesthesia on oxygen consumption: A meta-analysis guiding future studies on perioperative oxygen transport. Acta Anaesthesiol Scand. 2019;63(2):144–53.

5. Carsetti A, Amici M, Bernacconi T, Brancaleoni P, Cerutti E, Chiarello M, et al. Estimated oxygen extraction versus dynamic parameters of fluid-responsiveness for perioperative hemodynamic optimization of patients undergoing non-cardiac surgery: a non-inferiority randomized controlled trial. BMC anesthesiol. 2020;20(1):87.

6. Biro P. A formula to calculate oxygen uptake during low flow anesthesia based on FIO2 measurement. J Clin Monit Comput. 1998;14(2):141–4.

7. Leonard IE, Weitkamp B, Jones K, Aittomaki J, Myles PS. Measurement of systemic oxygen uptake during low-flow anaesthesia with a standard technique vs. a novel method. Anaesthesia. 2002;57(7):654–8.

8. McLellan S, Walsh TS. Oxygen delivery and haemoglobin. Cont Ed Anaesth Crit Care Pain 2004;4(4):123–6.

9. Keinanen O, Takala J. Calculated versus measured oxygen consumption during and after cardiac surgery. Is it possible to estimate lung oxygen consumption? Acta Anaesthesiol Scand. 1997;41(7):803–9.

10. Peyton PJ, Robinson GJ. Measured pulmonary oxygen consumption: difference between systemic oxygen uptake measured by the reverse Fick method and indirect calorimetry in cardiac surgery. Anaesthesia. 2005;60(2):146–50.

11. Saito H, Minamiya Y, Kawai H, Motoyama S, Katayose Y, Kimura K, et al. Estimation of pulmonary oxygen consumption in the early postoperative period after thoracic surgery. Anaesthesia. 2007;62(7):648–53.

12. Jakobsson J, Noren C, Hagel E, Kalman S, Bartha E. Peri-operative oxygen consumption revisited: An observational study in elderly patients undergoing major abdominal surgery. Eur J Anaesthesiol. 2021;38(1):4–12.

13. Wasserman K. Principles of exercise testing and interpretation including patophysiology and clinical applications. 5th ed. Philadelphia: Wolters Kluwer Health/Lippincott Williams & Wilkins; 2012.

14. Dunn JO MM, Grocott MP. Physiology of oxygen transport. BJA Ed. 2016;16(10):341–8.

15. Wang Y, Moss J, Thisted R. Predictors of body surface area. J Clin Anesth. 1992;4(1):4–10.

16. van Beest P, Wietasch G, Scheeren T, Spronk P, Kuiper M. Clinical review: use of venous oxygen saturations as a goal - a yet unfinished puzzle. Crit Care. 2011;15(5):232.

17. Pearse R, Dawson D, Fawcett J, Rhodes A, Grounds RM, Bennett ED. Changes in central venous saturation after major surgery, and association with outcome. Crit Care. 2005;9(6):R694–9.

18. Chemtob RA, Eskesen TG, Moeller-Soerensen H, Perner A, Ravn HB. Systematic review of the association of venous oxygenation and outcome in adult hospitalized patients. Acta Anaesthesiol Scand. 2016;60(10):1367–78.

19. Bizouarn P, Soulard D, Blanloeil Y, Guillet A, Goarin Y. Oxygen consumption after cardiac surgery--a comparison between calculation by Fick’s principle and measurement by indirect calorimetry. Intensive Care Med. 1992;18(4):206–9.

20. Hofland J, Tenbrinck R, Eggermont AM, Eijck CH, Gommers D, Erdmann W. Effects of simultaneous aortocaval occlusion on oxygen consumption in patients. Clin Physiol Funct Imaging. 2003;23(5):275–81.

21. Myles PS, Cui J. Using the Bland-Altman method to measure agreement with repeated measures. Br J Anaesth. 2007;99(3):309–11.

22. Inadomi C, Terao Y, Yamashita K, Fukusaki M, Takada M, Sumikawa K. Comparison of oxygen consumption calculated by Fick’s principle (using a central venous catheter) and measured by indirect calorimetry. J. 2008;22(2):163–6.

23. Burtman DTM, Stolze A, Genaamd Dengler SEK, Vonk ABA, Boer C. Minimally Invasive Determinations of Oxygen Delivery and Consumption in Cardiac Surgery: An Observational Study. J Cardiothorac Vasc Anesth. 2018;32(3):1266–72.

24. Soussi S, Vallee F, Roquet F, Bevilacqua V, Benyamina M, Ferry A, et al. Measurement of Oxygen Consumption Variations in Critically Ill Burns Patients: Are the Fick Method and Indirect Calorimetry Interchangeable? Shock. 2017;48(5):532–8.

25. Bland JM, Altman DG. Statistical methods for assessing agreement between two methods of clinical measurement. Lancet. 1986;1(8476):307–10.

26. Epstein CD, Peerless JR, Martin JE, Malangoni MA. Comparison of methods of measurements of oxygen consumption in mechanically ventilated patients with multiple trauma: the Fick method versus indirect calorimetry. Crit Care Med. 2000;28(5):1363–9.

27. Hanique G, Dugernier T, Laterre PF, Dougnac A, Roeseler J, Reynaert MS. Significance of pathologic oxygen supply dependency in critically ill patients: comparison between measured and calculated methods. Intensive Care Med. 1994;20(1):12–8.

28. Hofland J, Tenbrinck R, van Eijck CH, Eggermont AM, Gommers D, Erdmann W. Comparison of closed circuit and Fick-derived oxygen consumption in patients undergoing simultaneous aortocaval occlusion. Anaesthesia. 2003;58(4):377–84.

29. Myles PS, McRae R, Ryder I, Hunt JO, Buckland MR. Association between oxygen delivery and consumption in patients undergoing cardiac surgery. Is there supply dependence? Anaesth Intensive Care. 1996;24(6):651–7.

30. Critchley LA, Lee A, Ho AM. A critical review of the ability of continuous cardiac output monitors to measure trends in cardiac output. Anesth Analg. 2010;111(5):1180–92.

31. Liljequist D, Elfving B, Skavberg Roaldsen K. Intraclass correlation - A discussion and demonstration of basic features. PLoS ONE. 2019;14(7):e0219854.

32. Bland JM, Altman DG. Measuring agreement in method comparison studies. Stat Methods Med Res. 1999;8(2):135–60.

33. Bizouarn P, Blanloeil Y, Pinaud M. Comparison between oxygen consumption calculated by Fick’s principle using a continuous thermodilution technique and measured by indirect calorimetry. Br J Anaesth. 1995;75(6):719–23.

34. Smithies MN, Royston B, Makita K, Konieczko K, Nunn JF. Comparison of oxygen consumption measurements: indirect calorimetry versus the reversed Fick method. Crit Care Med. 1991;19(11):1401–6.

35. Stuart-Andrews C, Peyton P, Robinson G, Terry D, O’Connor B, Van der Herten C, et al. Non-invasive metabolic monitoring of patients under anaesthesia by continuous indirect calorimetry--an in vivo trial of a new method. Br J Anaesth. 2007;98(1):45–52.

36. Walsh TS, Hopton P, Lee A. A comparison between the Fick method and indirect calorimetry for determining oxygen consumption in patients with fulminant hepatic failure. Crit Care Med. 1998;26(7):1200–7.

37. Loer SA, Scheeren TW, Tarnow J. How much oxygen does the human lung consume? Anesthesiology. 1997;86(3):532–7.

38. Oudemans-van Straaten HM, Scheffer GJ, Eysman L, Wildevuur CR. Oxygen consumption after cardiopulmonary bypass--implications of different measuring methods. Intensive Care Med. 1993;19(2):105–10.

39. Schaffartzik W, Sanft C, Schaefer JH, Spies C. Different dosages of dobutamine in septic shock patients: determining oxygen consumption with a metabolic monitor integrated in a ventilator. Intensive Care Med. 2000;26(12):1740–6.

40. Jolliet P, Thorens JB, Nicod L, Pichard C, Kyle U, Chevrolet JC. Relationship between pulmonary oxygen consumption, lung inflammation, and calculated venous admixture in patients with acute lung injury. Intensive Care Med. 1996;22(4):277–85.

41. Brandi LS, Bertolini R, Pieri M, Giunta F, Calafa M. Comparison between cardiac output measured by thermodilution technique and calculated by O2 and modified CO2 Fick methods using a new metabolic monitor. Intensive Care Med. 1997;23(8):908–15.

42. Allingstrup MJ, Kondrup J, Perner A, Christensen PL, Jensen TH, Henneberg SW. Indirect Calorimetry in Mechanically Ventilated Patients: A Prospective, Randomized, Clinical Validation of 2 Devices Against a Gold Standard. JPEN J Parenter Enteral Nutr. 2017;41(8):1272–7.

43. Rehal MS, Fiskaare E, Tjader I, Norberg A, Rooyackers O, Wernerman J. Measuring energy expenditure in the intensive care unit: a comparison of indirect calorimetry by E-sCOVX and Quark RMR with Deltatrac II in mechanically ventilated critically ill patients. Crit Care. 2016;20:54.

44. Stuart-Andrews CR, Peyton P, Robinson GJ, Terry D, O’Connor B, Van der Herten C, et al. In vivo validation of the M-COVX metabolic monitor in patients under anaesthesia. Anaesth Intensive Care. 2007;35(3):398–405.

45. Lamia B, Kim HK, Severyn DA, Pinsky MR. Cross-comparisons of trending accuracies of continuous cardiac-output measurements: pulse contour analysis, bioreactance, and pulmonary-artery catheter. J Clin Monit Comput. 2018;32(1):33–43.

46. Hadian M, Kim HK, Severyn DA, Pinsky MR. Cross-comparison of cardiac output trending accuracy of LiDCO, PiCCO, FloTrac and pulmonary artery catheters. Crit Care. 2010;14(6):R212.

47. Beattie C, Moores C, Thomson AJ, Nimmo AF. The effect of anaesthesia and aortic clamping on cardiac output measurement using arterial pulse power analysis during aortic aneurysm repair. Anaesthesia. 2010;65(12):1194–9.

48. Lequeux PY, Bouckaert Y, Sekkat H, Van der Linden P, Stefanidis C, Huynh CH, et al. Continuous mixed venous and central venous oxygen saturation in cardiac surgery with cardiopulmonary bypass. Eur J Anaesthesiol. 2010;27(3):295–9.

49. van Beest PA, van Ingen J, Boerma EC, Holman ND, Groen H, Koopmans M, et al. No agreement of mixed venous and central venous saturation in sepsis, independent of sepsis origin. Crit Care. 2010;14(6):R219.

50. Soussi MS, Jebali MA, Le Manach Y, Nasri M, Zouari B, Chenik S, et al. Central venous saturation is not an alternative to mixed venous saturation during cardiopulmonary bypass in coronary artery surgery patients. Perfusion. 2012;27(4):300–6.

51. Reinhart K, Kuhn HJ, Hartog C, Bredle DL. Continuous central venous and pulmonary artery oxygen saturation monitoring in the critically ill. Intensive Care Med. 2004;30(8):1572–8.

52. el-Masry A, Mukhtar AM, el-Sherbeny AM, Fathy M, el-Meteini M. Comparison of central venous oxygen saturation and mixed venous oxygen saturation during liver transplantation. Anaesthesia. 2009;64(4):378–82.

53. Dueck MH, Klimek M, Appenrodt S, Weigand C, Boerner U. Trends but not individual values of central venous oxygen saturation agree with mixed venous oxygen saturation during varying hemodynamic conditions. Anesthesiology. 2005;103(2):249–57.

